# COVID-19 vaccines effectiveness against symptomatic SARS-CoV-2 during Delta variant surge: a population-based case-control study in St. Petersburg, Russia

**DOI:** 10.1101/2022.01.24.22269714

**Authors:** Anton Barchuk, Anna Bulina, Mikhail Cherkashin, Natalia Berezina, Tatyana Rakova, Darya Kuplevatskaya, Oksana Stanevich, Dmitriy Skougarevskiy, Artemiy Okhotin

## Abstract

**Background:** Studies of mRNA and vector-based vaccines used in different countries report acceptable levels of effectiveness against SARS-CoV-2 infection caused by the Delta variants of SARS-CoV-2. No studies estimated vaccine effectiveness (VE) of Gam-COVID-Vac and other vaccines used in Russia against symptomatic infection with Delta variant. In this population-based case-control study, we aimed to estimate the effectiveness of the Russian COVID-19 vaccines against symptomatic SARS-CoV-2 during the recent outbreak caused by the Delta VOC in October 2021 in St. Petersburg, Russia.

**Methods:** In a population-based case-control study with density sampling of controls, we acquired information on cases and controls from two independent studies conducted in St. Petersburg. Cases were symptomatic patients with confirmed SARS-CoV-2 (using polymerase chain reaction (PCR) test) referred to low-dose computed tomography (LDCT) triage in two outpatient centres between October 6 and 14, 2021 during the Delta variant outbreak. We recruited the controls during the representative survey of the seroprevalence study conducted during the same period in St. Petersburg using random digit dialling. In the primary analysis, we used logistic regression models to estimate the adjusted (age, gender, and history of confirmed COVID-19) VE against symptomatic SARS-CoV-2 resulted in a referral to triage centre for three vaccines used in Russia: Gam-COVID-Vac, EpiVacCorona, and CoviVac.

**Findings:** We included 1,254 cases and 2,747 controls recruited between the 6th and 14th of October in the final analysis. VE was 56% (95% CI: 48 to 63) for Gam-COVID-Vac (Sputnik V), 49% (95% CI: 29 to 63) for 1-dose Gam-COVID-Vac (Sputnik V) or Sputnik Light, -58% (95% CI: -225 to 23) for EpiVacCorona and 40% (95% CI: 3 to 63) for CoviVac. Without adjustment for the history of confirmed COVID-19 VE for all vaccines was lower, except for one-dose Gam-COVID-Vac (Sputnik Light). The adjusted VE was slightly lower in women — 51% (95% CI: 39 to 60) than men — 65% (95% CI: 5 to 73). It was also higher in younger age. However, in the analysis restricted to participants without a history of confirmed COVID-19, the differences in VE by age group were smaller.

**Interpretation:** In contrast to other Russian vaccines, Gam-COVID-Vac is effective against symptomatic SARS-CoV-2 infection caused by Delta VOC. Effectiveness is likely higher than the estimated 56% due to bias arising from high prevalence of the past COVID-19 in St. Petersburg.

**Funding:** Population-based survey in St. Petersburg was funded by Polymetal International, plc.

## Introduction

Mounting evidence suggests that vaccines remain effective against new variants of SARS-CoV-2, the virus that started the COVID-19 pandemic. Studies of mRNA and vector-based vaccines used in different countries report acceptable levels of effectiveness against SARS-CoV-2 infection caused by the new variants of concern (VOC) [1–7]. Waning immunity against new VOCs is another emerging concern as the follow-up time after the start of the vaccination programmes increases [8]. Therefore, timely and ongoing monitoring of the real-world effects for all vaccines used in programmes worldwide is crucial, given the geographical and temporal variation in global vaccine uptake and the spread of the new virus variants [9].

Only several international studies assessed the vaccine effectiveness (VE) of Gam-COVID-Vac [10], a vaccine dominating the Russian COVID-19 vaccination programme. A study conducted in Hungary provided data on comparative VE in a national population-based programme that used several vaccines, including Gam-COVID-Vac from Russia and HB02 from China [11]. Gam-COID-Vac effectiveness against SARS-CoV-2 infection and COVID-19-related mortality was comparable to mRNA vaccines and slightly superior to other vector-based vaccines. However, the Hungary study covers the period before the spread of the Delta VOC. The case-control study in St. Petersburg was the first available evidence of vaccine effectiveness (VE) against referral to hospital in patients with symptomatic SARS-CoV-2 infection in Russia during the spread of the Delta VOC [7]. However, that study had several limitations. The information on the vaccine type was not available, so the estimated VE represented an average effect of three vaccines used in St. Petersburg: Gam-COVID-Vac, EpiVacCorona [12], and CoviVac [13]. It was safe to assume that the St. Petersburg study approximated the effectiveness of Gam-COVID-Vac given that it accounted for 95% of city vaccinations during the period under study. However, the VE for the two other vaccines used in Russia — EpiVacCorona and CoviVac — remained unclear. More importantly, that study did not provide direct evidence on the VE against SARS-CoV-2 infection and symptomatic disease.

Another point of concern in observational studies of VE is the proportion of individuals with natural immunity which protects from re-infection [14, 15]. If case-control studies include individuals with immunity after infection as controls, that will likely underestimate the actual VE against SARS-CoV-2. To illustrate this point, more than 45% of the population have contracted the SARS-CoV-2 by the end of April, 2021 in St. Petersburg, Russia [16]. Preliminary study results show that seroprevalence in unvaccinated may be more than 75% in October, 2021 [17].

In this population-based case-control study, we aimed to estimate the effectiveness of the Russian COVID-19 vaccines against symptomatic SARS-CoV-2 during the recent outbreak caused by the Delta VOC in October 2021 in St. Petersburg, Russia.

## Methods

### Population and study design

This report summarised the results of the population-based case-control study of VE against the symptomatic disease with density sampling of controls conducted in October 2021 in St. Petersburg, Russia. We acquired information on cases and controls from two ongoing independent studies in St. Petersburg [7, 16] with population-based controls sampled at similar points in time when cases have occurred.

Our cases were symptomatic patients with confirmed SARS-CoV-2 (using polymerase chain reaction (PCR) test) referred to low-dose computed tomography (LDCT) triage. Our previous report describes in detail the source and information we collected for the cases [7]. In brief, we collected individual-level data from two outpatient triage centres of the Medical Institute named after Berezin Sergey (MIBS), a private medical facility contracted by the city government to provide triage service for nearly half of the city districts. Triage centres continuously collected this information from August 2021. In addition to the data collected for our previous case-control study, we added information on the vaccine type (Gam-COVID-Vac (Sputnik V and Sputnik Light), EpiVacCorona, and CoviVac) and the history of confirmed COVID-19, which we defined as the positive PCR test at least two months before the current episode.

We recruited controls from a survey of the seroprevalence study conducted in the same period in St. Petersburg. Our previous reports described the serosurvey design in detail [16, 18]. It includes a two-step approach: a survey conducted using random digit dial (RDD) and computer-assisted telephone interview (CATI) followed by an invitation to a serological test. The survey samples were representative of the population of the city in terms of sociodemographic characteristics. In addition, CATI included questions related to history of confirmed COVID-19 and vaccination status in line with information collected from cases.

For this study, we extracted information on all SARS-CoV-2 patients referred to the triage centre between the 6th and 14th of October, 2021 (which was the period when controls data from the survey were available).

### Vaccination status, outcomes, and other variables

Vaccination status in our study was self-reported. Both cases and controls were asked about their vaccination status, vaccine type, the number of doses, and dates for the doses. If the exact day was not reported, at least the month and the year of vaccination were collected. Three vaccines were available in St. Petersburg during the pandemic: Gam-COVID-Vac two-dose (Sputnik V) and one-dose regimen (Sputnik Light), EpiVacCorona and CoviVac (both two-dose regimens). All vaccines were approved for primary vaccination, but Sputnik Light was specifically recommended as preferred option for the booster after COVID-19 infection. We assigned the full vaccination status to all participants (both cases and controls) who reported the second dose in September 2021 and earlier. We set full vaccination status for participants with uncertain vaccination dates, but this assumption was further addressed in the sensitivity analysis. We assigned partial vaccination status to participants who received only one dose but did not satisfy the criteria for full vaccination status. We have also analysed Sputnik Light vaccines as a distinct group. Participants who received one dose of Gam-COVID-Vac, but had not received the second dose till October entered the Sputnik Light group.

The primary outcome was the referral to LDCT triage with symptomatic infection and the positive PCR for SARS-CoV-2. Cases were patients referred to LDCT triage who underwent brief physical examination and computed tomography (CT). We were collecting CT-score (five gradations from 0 to 4), which represents lung segment involvement used in our previous study [7]. The secondary outcome was any lung injury as reported by LDCT in the triage centre (CT-score 1, 2, 3, or 4). We did not collect or use the hospital referral as an outcome in contrast to our previous study because official criteria for hospitalisation changed in the autumn of 2021 in St. Petersburg. Other variables include age, gender, history of confirmed COVID-19.

### Analysis

We modelled our study plan following the WHO interim guidance to evaluate COVID-19 vaccine effectiveness [19]. We used unconditional logistic regression for our primary and secondary outcomes to estimate odds ratios (ORs) for vaccination status among cases and controls, which approximates ORs for the outcomes among the vaccinated and non-vaccinated patients. In the presence of a density control sampling scheme, it approximates the rate ratio from the respective cohort study [20]. This study design was previously used to assess risk factors for respiratory infections [21].

The VE was calculated as 100% *×* (1 *− OR*) adjusted for age and gender. Sample size of 1,198 cases and 2,747 controls, and 1,175 patients with the complete vaccination status (exposure level of 29.8% for Sputnik V) provides 80% power to detect an odds ratio of 0.80 (or the VE of 20%) at the 5% alpha level. For 243 fully vaccinated with one dose Sputnik Light (exposure level of 6.2%) odds ratio of 0.63 (or VE of 37%) is detectable. For 104 fully vaccinated with CoviVac (exposure level of 2.6%) odds ratio of 0.45 (or VE of 55%) is detectable. However, for 28 fully vaccinated with EpiVacCorona (exposure level of 0.7%) odds ratio of only 0.03 (or VE of only 97%) is detectable.

We corrected VE for the history of confirmed COVID-19 infection by performing the analyses in the dataset restricted to participants without the history of confirmed COVID-19.

In the sensitivity analysis, we explored the possible extent of misclassification. To address the misclassification for the vaccination dates for some participants, we have changed the status of controls with uncertain dates of vaccination to non-vaccinated. In addition, in another sensitivity analysis, we assumed that cases with missing vaccine names received Gam-COVID-Vac. We reported all results of sensitivity analysis in Supplementary Materials.

### Ethical considerations

The Ethics Committee of the MIBS approved the VE study on June 21, 2021. The Ethics Committee of the Pavlov First Saint Petersburg State Medical University approved the joint study of COVID-19 VE in St. Petersburg on July 15, 2021. All research was performed following the relevant guidelines and regulations. All participants signed the informed consent upon referral to the LDCT triage. The joint study of COVID-19 vaccine effectiveness in St. Petersburg was registered at ClinicalTrials.gov (NCT04981405, date of registration: August 4, 2021). This publication covers the data collected at the two outpatient centres of the MIBS that contributed to the study data. The Research Planning Board of the European University at St. Petersburg and the Ethics Committee of the Clinic “Scandinavia” approved the seroprevalence study on May 20, 2020 and May 26, 2020, respectively. Consent was obtained from all participants of the study. The study was registered with the following identifiers:Clinicaltrials.gov (NCT04406038, submitted on May 26, 2020, date of registration: May 28, 2020) and ISRCTN registry (ISRCTN11060415, submitted on May 26, 2020, date of registration: May 28, 2020).

### Data sharing

All analyses were conducted in R, study data and code is available online (https://github.com/eusporg/spb_covid_study20).

### Contribution and role of the funding source

Polymetal International plc funded the serological study. The main funder had no role in study design, data collection, data analysis, data interpretation, report writing, or decision to submit the publication. The European University at St. Petersburg and MIBS had access to the study data. The European University at St. Petersburg had the final responsibility to submit for publication.

## Results

Overall, 1,254 cases and 2,747 controls were included in the final analysis. Study participants characteristics are presented in Table 1. Before October 2021, among cases, 288 (23.0%) received Gam-COVID-Vac (Sputnik V), 57 (4.5%) — one dose Sputnik Light, 19 (1.5%) — EpiVacCorona, and 37 (3.0%) — CoviVac. Similar proportions among controls were 937 (34.1%), 214 (7.8%), 15 (0.5%), and 88 (3.2%), respectively. Cases were on average older (27.9% were older than 60 years compared to 19.1% among controls) and the proportion of women was also higher among cases (63.1% among cases vs 53.7% among controls). Only 2 (0.2%) patients who were referred for LDCT triage had documented past COVID-19 confirmed by PCR test. The proportion of controls who reported positive PCR test in the past was 22.1%. Both cases and controls were relatively uniformly distributed across recruitment dates (October 6–14, 2021).

**Table 1.**
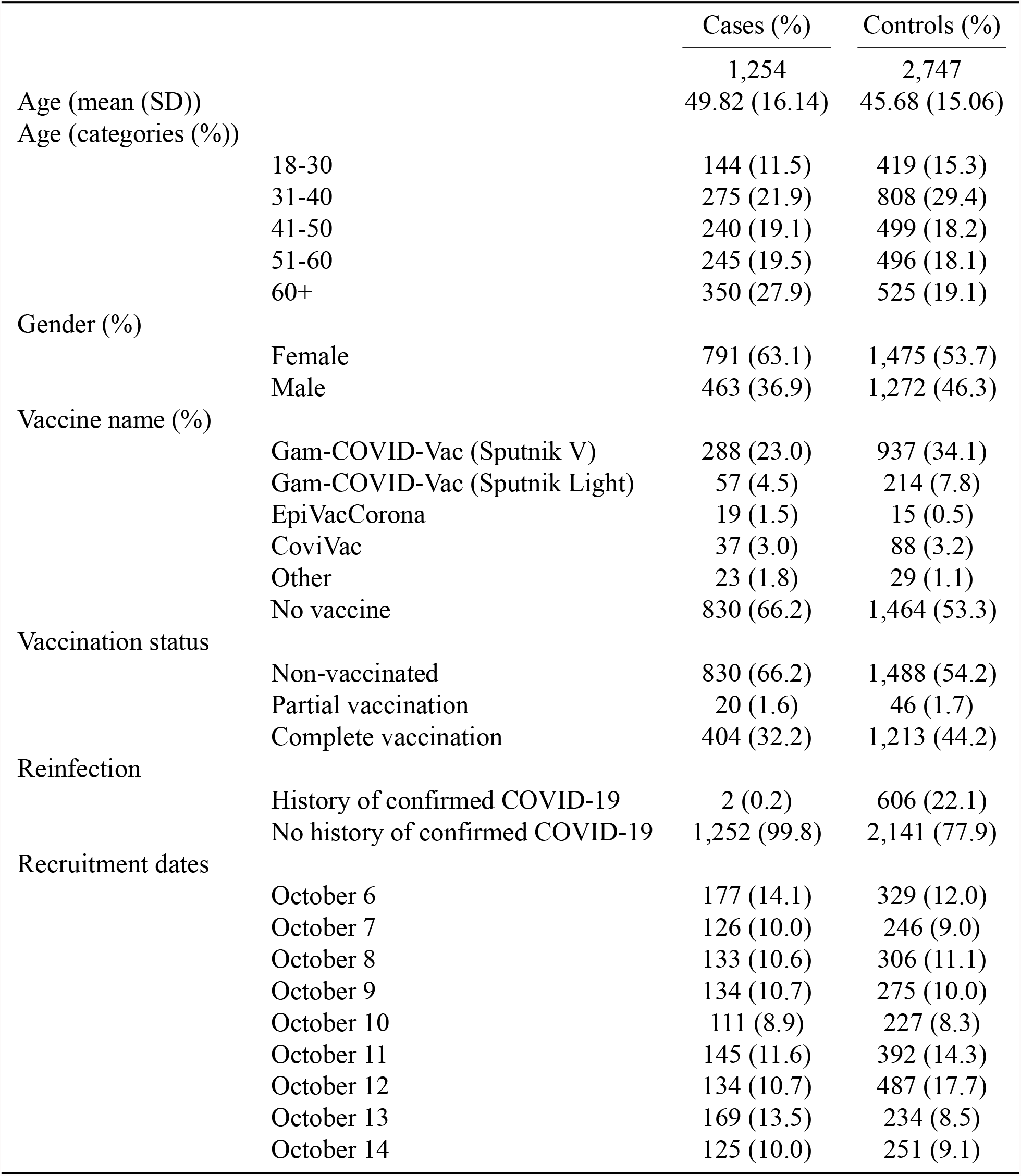
Characteristics of Cases and Controls

|  | Cases (%) | Controls (%) |
| --- | --- | --- |
|  | 1,254 | 2,747 |
| Age (mean (SD)) | 49.82 (16.14) | 45.68 (15.06) |
| Age (categories (%)) |  |  |
| 18-30 | 144 (11.5) | 419 (15.3) |
| 31-40 | 275 (21.9) | 808 (29.4) |
| 41-50 | 240 (19.1) | 499 (18.2) |
| 51-60 | 245 (19.5) | 496 (18.1) |
| 60+ | 350 (27.9) | 525 (19.1) |
| Gender (%) |  |  |
| Female | 791 (63.1) | 1,475 (53.7) |
| Male | 463 (36.9) | 1,272 (46.3) |
| Vaccine name (%) |  |  |
| Gam-COVID-Vac (Sputnik V) | 288 (23.0) | 937 (34.1) |
| Gam-COVID-Vac (Sputnik Light) | 57 (4.5) | 214 (7.8) |
| EpiVacCorona | 19 (1.5) | 15 (0.5) |
| CoviVac | 37 (3.0) | 88 (3.2) |
| Other | 23 (1.8) | 29 (1.1) |
| No vaccine | 830 (66.2) | 1,464 (53.3) |
| Vaccination status |  |  |
| Non-vaccinated | 830 (66.2) | 1,488 (54.2) |
| Partial vaccination | 20 (1.6) | 46 (1.7) |
| Complete vaccination | 404 (32.2) | 1,213 (44.2) |
| Reinfection |  |  |
| History of confirmed COVID-19 | 2 (0.2) | 606 (22.1) |
| No history of confirmed COVID-19 | 1,252 (99.8) | 2,141 (77.9) |
| Recruitment dates |  |  |
| October 6 | 177 (14.1) | 329 (12.0) |
| October 7 | 126 (10.0) | 246 (9.0) |
| October 8 | 133 (10.6) | 306 (11.1) |
| October 9 | 134 (10.7) | 275 (10.0) |
| October 10 | 111 (8.9) | 227 (8.3) |
| October 11 | 145 (11.6) | 392 (14.3) |
| October 12 | 134 (10.7) | 487 (17.7) |
| October 13 | 169 (13.5) | 234 (8.5) |
| October 14 | 125 (10.0) | 251 (9.1) |

In the primary analysis without accounting for the history of confirmed COVID-19, the VE against symptomatic PCR-confirmed SARS-CoV-2 infection adjusted for age and gender was 49% (95% CI: 40–56) for Gam-COVID-Vac (Sputnik V), 51% (95% CI: 32–64) for 1-dose Gam-COVID-Vac (Sputnik V) or Sputnik Light, -84% (95% CI: -267–8)) for EpiVacCorona, and 35% (95% CI: -3–59) for CoviVac. In the analysis restricted to participants without a history of confirmed COVID-19, all point estimates of VE moved upwards, except for the Sputnik Light group. The VE was 56% (95% CI: 48–63) for Gam-COVID-Vac (Sputnik V), 49% (95% CI: 29–63) for 1-dose Gam-COVID-Vac (Sputnik V) or Sputnik Light, -58% (95% CI: -225–23)) for EpiVacCorona, and 40% (95% CI: 3–63) for CoviVac (Table 2).

**Table 2.** Effectiveness of Vaccination against Symptomatic PCR-confirmed SARS-CoV-2

|  | Crude VE<br>(95% CI) | VE adjusted for<br>age and gender<br>(95% CI) | VE adjusted for age, gender, and<br>history of confirmed COVID-19<br>(95% CI) |
| --- | --- | --- | --- |
| Gam-COVID-Vac (2-dose Sputnik V) | 43% (34–52) | 49% (40–56) | 56% (48–63) |
| Gam-COVID-Vac (1-dose Sputnik Light) | 49% (31–63) | 51% (32–64) | 49% (29–63) |
| EpiVacCorona | -105% (-322–1) | -84% (-267–8) | -58% (-225–23) |
| CoviVac | 34% (-3–49) | 35% (-3–59) | 40% (3–63) |
| Other | -22% (-127–34) | -26% (-125–29) | -1% (-82–44) |
| Any vaccine: partial vaccination | 22% (-33–54) | 24% (-33–56) | 25% (-33–58) |

Crude and adjusted VE against any lung injury following the LDCT assessment is presented in Table 3. In the analysis restricted to participants without a history of confirmed COVID-19, all point estimates of VE against lung injury also moved upwards.

**Table 3.** Effectiveness of Vaccination against Any Lung Injury

|  | Crude VE<br>(95% CI) | VE adjusted for<br>age and gender<br>(95% CI) | VE adjusted for age, gender, and<br>history of confirmed COVID-19<br>(95% CI) |
| --- | --- | --- | --- |
| Gam-COVID-Vac (2-dose Sputnik V) | 57% (48–64) | 64% (56–70) | 69% (62–74) |
| Gam-COVID-Vac (1-dose Sputnik Light) | 56% (36–70) | 58% (38–71) | 58% (37–72) |
| EpiVacCorona | -82% (-280–13) | -52% (-205–24) | -30% (-165–37) |
| CoviVac | 43% (4–66) | 44% (5–67) | 49% (11–70) |
| Other | -22% (-137–37) | -24% (-130–33) | -1% (-89–46) |
| Any vaccine: partial vaccination | 33% (-24–66) | 35% (-30–64) | 36% (-25–67) |

The adjusted VE against symptomatic SARS-CoV-2 infection was slightly lower in women (51%, 95% CI: 39–60) than men (65%, 95% CI: 53–73). It was also higher in younger age (Table 4). However, in the analysis restricted to participants without a history of confirmed COVID-19, the differences in VE by age group were smaller.

**Table 4.** Effectiveness of Vaccination Against Symptomatic PCR-confirmed SARS-CoV-2 by Age and Gender

|  | Crude VE (95% CI) | VE adjusted for confirmed COVID-19 history (95% CI) |
| --- | --- | --- |
| Age groups |  |  |
| 18–30 | 70% (47–83) | 73% (51–85) |
| 31–40 | 54% (34–68) | 57% (38–71) |
| 41–50 | 53% (32–69) | 58% (38–72) |
| 51–60 | 39% (14–56) | 51% (30–65) |
| 60+ | 41% (21–56) | 52% (35–65) |
| Gender |  |  |
| Male | 59% (47–68) | 65% (53–73) |
| Female | 42% (29–52) | 51% (39–60) |

In the sensitivity analysis, misclassification related to vaccine name and vaccination dates did not dramatically change VE estimates.

## Discussion

This is the first study examining the comparative effectiveness of three COVID-19 vaccines used in Russia. Our study results assure that Gam-COVID-Vac is highly effective against symptomatic SARS-CoV-2 and severe COVID-19 pneumonia during the Delta VOC spread. We have shown that Gam-COVID-Vac provides at least 58% protection against symptomatic infection caused by the Delta VOC. However, the effectiveness is likely to be higher as it is difficult to account for all past SARS-CoV-2 infections in the Russian population. Furthermore, past COVID-19 is associated with decreased vaccine uptake in Russia. Our study’s apparent strength is the attempt to account for past infections in both cases and controls and show the resulting direction of possible bias. Assuming natural immunity is protective against re-infection, failure to account for it in populations with high seroprevalence would bias VE estimates downwards. Another possible representation of this is the change in Gam-COVID-Vac VE by age group after accounting for the history of confirmed COVID-19.

We have shown that the bias related to a significant number of unvaccinated individuals with a history of COVID-19 will likely lead to underestimating the effectiveness of vaccination from observational data. However, we remain uncertain about the possible magnitude of this bias. The individual-level data on past asymptomatic infection is challenging to obtain, if possible at all. Preliminary results show that seroprevalence in unvaccinated may be more than 75% in October, 2021 [17] in St. Petersburg. In countries with higher vaccine uptake and lower seroprevalence VE estimated in observational studies is higher [5], but these estimates are not directly comparable with our results. The Omicron VOC spread will likely make the interpretation of the VE studies even more difficult. Direct comparison similar to the Hungarian study will be needed for the new variants [11].

In contrast to Gam-COVID-Vac, two other vaccines, EpiVacCorona and CoviVac, were not similarly effective against symptomatic infection caused by Delta VOC of SARS-CoV-2 in our study. Both vaccines were relatively rare in the population of St. Petersburg, and our study was underpowered for them. However, our study provides reasonable doubts about possible effectiveness of EpiVacCorona against new emerging VOCs. CoviVac usefulness is also doubtful in the presence of highly effective Gam-COVID-Vac. More studies are needed to assess the VE of EpiVacCorona and CoviVac against new variants of SARS-CoV-2 for them to be used in the ongoing vaccination programme. Unfortunately, efficacy data is currently available only for Gam-COVID-Vac [22]. Booster campaigns that are now gaining more scientific support should only utilise vaccines with proven efficacy and effectiveness [23–25].

It is also worth mentioning that while the VE for CoviVac was beyond the VE for Gam-COVID-Vac, the estimate for EpiVacCorona VE was negative. The efficacy is not likely to be negative, so our results have two realistic explanations. First, individuals could change their behaviour after vaccination, but more likely negative VE is a marker for the bias arising from the undercounting of past COVID-19 in controls.

This is our second VE study in St. Petersburg, Russia, and it provides a promising independent and timely framework for assessing COVID-19 vaccines in Russia. Population-based case-control studies represent a critical post-registration tool to monitor VE against emerging SARS-CoV-2 VOCs. The Omicron VOC pandemic has not involved Russia by the end of 2021, but there are few doubts that it will affect the course of the pandemic in Russia as previously the Delta VOC had [16]. The lack of real-world evidence may be one of the reasons behind the modest uptake of vaccination in Russia. The majority in Russia does not deny the idea of vaccination but is hesitant [26].

Despite the wide use of case-control studies to assess VE, researchers should be aware of all possible biases arising from this study design. Unfortunately, the golden standard to estimate VE — randomised trials — are not applicable in the rapidly changing epidemiological situation, and we have to rely on observational study design. The varying VE against different SARS-CoV-2 variants is an example of a lack of generalizability for the results of randomised trials. Our study underlines the biases related to the population under study, but additional biases arise from the misclassification of exposure, e.g., vaccination status [27]. The self-reported vaccination status is an important limitation of our study. Several survey participants included in the control group have not reported the exact date of vaccination. While the overall number of such individuals was low, we assumed that the vaccination date for such individuals is likely to be several months from the interview date. However, we assigned them a “non-vaccinated” status in our sensitivity analysis, and the estimates were only slightly affected. Our definition for full vaccination status was also very conservative, as we decided to accept a minimum of six days between the second vaccine dose and study inclusion. While our decision was driven by the idea that we should not exclude participants without an exact date of vaccination, we do not think that this assumption would significantly bias the results. However, most of the studies choose 14-day period [5], and that should be taken into account when comparing our results to other studies.

We have undertaken additional attempts to identify cases (patients with symptomatic SARS-CoV-2 in October, 2021) who had the history of confirmed COVID-19 more than two months before the current episode. We were able to identify only two cases of re-infection. While underreporting may occur, it is also likely that a patient with re-infection that requires additional diagnostic follow-up is an infrequent event. Absolute risks of re-infection, especially of severe disease, are low for the Alpha, Beta, and Delta VOCs [28, 29]. However, more studies are needed to observe the risk of re-infection with new Omicron VOCs, as it is likely to be higher [30]. Overall, the risks may still be lower in absolute terms than for primary infection.

In our study, the VE in the Sputnik Light group was similar to two-dose Gam-COVID-Vac. However, the correction for the history of confirmed COVID-19 did not move the Sputnik Light VE upwards. Single Gam-COVID-Vac vaccination labelled as Sputnik Light was used as a booster after the COVID-19, so it is likely that the prevalence of past COVID-19 is higher is this group. The VE for Sputnik Light could represent the combination of single-dose boosted natural immunity mixed with single-dose vaccine.

Some of these limitations are inherent to observational study design. Still, other difficulties can be overcome by establishing a pre-existing framework for real-time assessment of vaccine effectiveness as a part of epidemiological surveillance.

In conclusion, Gam-COVID-Vac effectiveness against symptomatic SARS-CoV-2 infection caused by Delta VOC is at least 56%, but is likely to be higher. However, estimating effectiveness is difficult due to the high prevalence of natural immunity in the population. Nevertheless, Gam-COVID-Vac significantly outperforms two other Russian vaccines whose effectiveness against symptomatic SARS-CoV-2 infection caused by Delta VOC is yet to be shown.

## Data Availability

Study data and code is available online.

https://github.com/eusporg/spb_covid_study20

## Authors’ contributions

Anton Barchuk, Artemy Okhotin, and Mikhail Cherkashin conceived the study. Anton Barchuk, Artemy Okhotin, Mikhail Cherkashin, and Oksana Stanevich coordinated the study. Anton Barchuk, Artemy Okhotin, Dmitriy Skougarevskiy, and Anna Bulina drafted the first version of the manuscript. Mikhail Cherkashin, Natalia Berezina, Tatyana Rakova, Darya Kuplevatskaya, and Oksana Stanevich contributed to drafting sections of the manuscript. Mikhail Cherkashin, Natalia Berezina, Tatyana Rakova, Darya Kuplevatskaya collected data for analyses. Anna Bulina and Anton Barchuk did data analyses. All authors participated in the study design, helped draft the manuscript, contributed to the interpretation of data, and read and approved the final manuscript.

## Declaration of interests

Anton Barchuk reports personal fees from AstraZeneca, MSD, and Biocad outside the submitted work. Other authors have no conflict of interest to declare.

## Acknowledgements

We acknowledge personal support from Vitaly Nesis (Chief Executive Officer, Polymetal International, plc). We are grateful to Alla Samoletova and Alexandra Vasilieva (European University at St. Petersburg) for science communication and administrative support. We thank the interviewers, nurses, and other personnel of the MIBS. We appreciate Yulia Stepantsova (Chursina) input in coordinating phone-based interviews, Daria Danilenko, Alexei Kouprianov and Daniel Munblit for insightful discussions. We also thank all study participants.

## Supplementary materials

**Table A1.** Characteristics of Cases and Controls by Vaccination Status. Sensitivity Analysis

|  |  | Overall (%) | Cases (%) | Controls (%) |
| --- | --- | --- | --- | --- |
| Missing vaccine names were assumed to be Gam-COVID-Vac |  |  |  |  |
| Vaccine name | Gam-COVID-Vac (2-dose Sputnik V) | 1,242 (31.0) | 305 (24.3) | 937 (34.1) |
|  | Gam-COVID-Vac (1-dose Sputnik Light) | 277 (6.9) | 63 (5.0) | 214 (7.8) |
|  | EpiVacCorona | 34 (0.8) | 19 (1.5) | 15 (0.5) |
|  | CoviVac | 125 (3.1) | 37 (3.0) | 88 (3.2) |
|  | Other | 29 (0.7) | 0 (0.0) | 29 (1.1) |
|  | No vaccine | 2,294 (57.3) | 830 (66.2) | 1,464 (53.3) |
| Vaccination status | Non-vaccinated | 2,318 (57.9) | 830 (66.2) | 1,488 (54.2) |
|  | Partial vaccination | 60 (1.5) | 14 (1.1) | 46 (1.7) |
|  | Complete vaccination | 1,623 (40.6) | 410 (32.7) | 1,213 (44.2) |
| Missing vaccine dates were assigned non-vaccination status |  |  |  |  |
| Vaccine name | Gam-COVID-Vac (2-dose Sputnik V) | 1,2225 (30.6) | 288 (23.0) | 937 (34.1) |
|  | Gam-COVID-Vac (1-dose Sputnik Light) | 271 (6.8) | 57 (4.5) | 214 (7.8) |
|  | EpiVacCorona | 34 (0.8) | 19 (1.5) | 15 (0.5) |
|  | CoviVac | 125 (3.1) | 37 (3.0) | 88 (3.2) |
|  | Other | 52 (1.3) | 23 (1.8) | 29 (1.1) |
|  | No vaccine | 2,294 (57.3) | 830 (66.2) | 1,464 (53.3) |
| Vaccination status | Non-vaccinated | 2,339 (58.5) | 831 (66.3) | 1,508 (54.9) |
|  | Partial vaccination | 66 (1.6) | 20 (1.6) | 46 (1.7) |
|  | Complete vaccination | 1,596 (39.9) | 403 (32.1) | 1,193 (43.4) |

**Table A2.** Effectiveness of Vaccination against Symptomatic PCR-confirmed SARS-CoV-2. Sensitivity Analysis

|  |  | Crude OR (95% CI) | Crude VE (95% CI) | OR adjusted<br>for age and gender (95% CI) | VE adjusted<br>for age and gender (95% CI) |
| --- | --- | --- | --- | --- | --- |
| Missing vaccine names were assumed to be Gam-COVID-Vac |  |  |  |  |  |
| Without correcting for the history of confirmed COVID-19 | Gam-COVID-Vac (2-dose Sputnik V) | 0.60 (0.51–0.70) | 40% (30–49) | 0.54 (0.47–0.64) | 46% (36–53) |
|  | Gam-COVID-Vac (1-dose Sputnik Light) | 0.59 (0.43–0.79) | 41% (21–57) | 0.56 (0.42–0.76) | 44% (24–58) |
|  | EpiVacCorona | 2.05 (0.99–4.22) | -105% (-322–0) | 1.85 (0.93–3.68) | -85% (-268–7) |
|  | CoviVac | 0.66 (0.42–1.03) | 34% (-3–58) | 0.65 (0.41–1.03) | 35% (-3–59) |
|  | Any vaccine: partial vaccination | 0.55 (0.30–1.00) | 45% (0–70) | 0.53 (0.29–1.00) | 47% (0–71) |
| After correcting for the history of confirmed COVID-19 | Gam-COVID-Vac (2-dose Sputnik V) | 0.52 (0.44–0.61) | 48% (39–56) | 0.47 (0.40–0.55) | 53% (45–60) |
|  | Gam-COVID-Vac (1-dose Sputnik Light) | 0.60 (0.43–0.82) | 40% (18–57) | 0.57 (0.41–0.79) | 43% (21–59) |
|  | EpiVacCorona | 1.77 (0.83–3.76) | -77% (-276–17) | 1.58 (0.77–2.26) | -58% (-225–23) |
|  | CoviVac | 0.62 (0.39–0.98) | 38% (2–61) | 0.60 (0.37–0.97) | 40% (3–63) |
|  | Any vaccine: partial vaccination | 0.53 (0.28–0.99) | 47% (1–72) | 0.52 (0.27–0.99) | 48% (1–73) |
| Missing vaccine dates were assigned non-vaccination status |  |  |  |  |  |
| Without correcting for the history of confirmed COVID-19 | Gam-COVID-Vac (2-dose Sputnik V) | 0.58 (0.49–0.68) | 42% (32–51) | 0.52 (0.45–0.62) | 48% (38–55) |
|  | Gam-COVID-Vac (1-dose Sputnik Light) | 0.54 (0.40–0.74) | 46% (26–60) | 0.52 (0.38–0.72) | 48% (29–62) |
|  | EpiVacVorona | 2.07 (1.00–4.27) | -107% (-327– -1) | 1.87 (0.94–3.72) | -87% (-272–6) |
|  | CoviVac | 0.71 (0.45–1.10) | 29% (-10–55) | 0.69 (0.44–1.10) | 31% (-10–56) |
|  | Other | 1.34 (0.71–2.52) | -34% (-152–29) | 1.36 (0.75–2.46) | -36% (-146–25) |
|  | Any vaccine: partial vaccination | 0.79 (0.46–1.34) | 21% (-34–54) | 0.78 (0.45–1.35) | 22% (-35–55) |
| After correcting for the history of confirmed COVID-19 | Gam-COVID-Vac (2-dose Sputnik V) | 0.50 (0.43–0.59) | 50% (41–57) | 0.45 (0.38–0.53) | 55% (47–62) |
|  | Gam-COVID-Vac (1-dose Sputnik Light) | 0.56 (0.40–0.77) | 44% (23–60) | 0.53 (0.38–0.74) | 47% (26–62) |
|  | EpiVacCorona | 1.80 (0.85–3.82) | -80% (-282–15) | 1.61 (0.78–3.30) | -61% (-230–22) |
|  | CoviVac | 0.67 (0.42–1.07) | 33% (-7–58) | 0.65 (0.40–1.06) | 35% (-6–60) |
|  | Other | 1.09 (0.57–2.08) | -9% (-108–43) | 1.10 (0.60–2.01) | -10% (-101–40) |
|  | Any vaccine: partial vaccination | 0.77 (0.44–1.34) | 23% (-34–56) | 0.76 (0.42–1.35) | 24% (-35–58) |

**Table A3.** Effectiveness of Vaccination against Lung Injury. Sensitivity Analysis

|  |  | Crude OR (95% CI) | Crude VE (95% CI) | OR adjusted<br>for age and gender (95% CI) | VE adjusted<br>for age and gender (95% CI) |
| --- | --- | --- | --- | --- | --- |
| Missing vaccine names were assumed to be Gam-COVID-Vac |  |  |  |  |  |
| Without correcting for the history of confirmed COVID-19 | Gam-COVID-Vac (2-dose Sputnik V) | 0.46 (0.39–0.56) | 54% (44–61) | 0.39 (0.32–0.47) | 61% (53–68) |
|  | Gam-COVID-Vac (1-dose Sputnik Light) | 0.49 (0.34–0.69) | 51% (31–66) | 0.47 (0.32–0.67) | 53% (33–68) |
|  | EpiVacCorona | 1.82 (0.87–3.80) | -82% (-280–13) | 1.52 (0.76–3.06) | -52% (-205–24) |
|  | CoviVac | 0.57 (0.34–0.96) | 43% (-4–66) | 0.56 (0.33–0.95) | 44% (5–67) |
|  | Any vaccine: partial vaccination | 0.48 (0.24–0.99) | 52% (1–76) | 0.46 (0.22–0.98) | 54% (2–78) |
| After correcting for the history of confirmed COVID-19 | Gam-COVID-Vac (2-dose Sputnik V) | 0.41 (0.34–0.50) | 59% (50–66) | 0.33 (0.28–0.41) | 67% (59–72) |
|  | Gam-COVID-Vac (1-dose Sputnik Light) | 0.49 (0.34–0.71) | 51% (29–66) | 0.47 (0.32–0.68) | 53% (32–68) |
|  | EpiVacCorona | 1.58 (0.74–3.36) | -58% (-236–26) | 1.30 (0.64–2.66) | -30% (-166–36) |
|  | CoviVac | 0.54 (0.32–0.92) | 46% (8–68) | 0.52 (0.30–0.89) | 48% (11–70) |
|  | Any vaccine: partial vaccination | 0.47 (0.23–0.98) | 53% (2–77) | 0.46 (0.21–1.00) | 54% (0–79) |
| Missing vaccine dates were assigned non-vaccination status |  |  |  |  |  |
| Without correcting for the history of confirmed COVID-19 | Gam-COVID-Vac (2-dose Sputnik V) | 0.44 (0.36–0.53) | 56% (47–64) | 0.37 (0.30–0.44) | 63% (56–70) |
|  | Gam-COVID-Vac (1-dose Sputnik Light) | 0.45 (0.31–0.65) | 56% (35–69) | 0.43 (0.30–0.63) | 57% (36–70) |
|  | EpiVacVorona | 1.84 (0.88–3.84) | -84% (-284–12) | 1.54 (0.77–3.09) | -54% (-209–23) |
|  | CoviVac | 0.61 (0.36–1.02) | 39% (-2–64) | 0.59 (0.35–1.09) | 41% (-1–65) |
|  | Other | 1.33 (0.68–2.59) | -33% (-159–32) | 1.31 (0.70–2.46) | -31% (-146–30) |
|  | Any vaccine: partial vaccination | 0.68 (0.37–1.25) | 32% (-25–63) | 0.66 (0.34–1.25) | 34% (-25–66) |
| After correcting for the history of confirmed COVID-19 | Gam-COVID-Vac (2-dose Sputnik V) | 0.39 (0.32–0.47) | 61% (53–68) | 0.32 (0.26–0.39) | 68% (61–74) |
|  | Gam-COVID-Vac (1-dose Sputnik Light) | 0.45 (0.31–0.67) | 55% (33–69) | 0.44 (0.29–0.65) | 56% (35–71) |
|  | EpiVaccorona | 1.60 (0.75–3.40) | -60% (-240–25) | 1.32 (0.64–2.69) | -32% (-169–36) |
|  | CoviVac | 0.58 (0.34–0.99) | 42% (1–66) | 0.55 (0.32–0.96) | 45% (4–68) |
|  | Other | 1.11 (0.56–2.18) | -11% (-118–44) | 1.07 (0.57–2.03) | -7% (-103–43) |
|  | Any vaccine: partial vaccination | 0.66 (0.35–1.24) | 34% (-24–65) | 0.65 (0.33–1.27) | 35% (-27–67) |

## Notes

### Author Declarations

The Ethics Committee of the MIBS approved the VE study on June 21, 2021. The Ethics Committee of the Pavlov First Saint Petersburg State Medical University approved the joint study of COVID-19 VE in St. Petersburg on July 15, 2021. All research was performed following the relevant guidelines and regulations. All participants signed the informed consent upon referral to the LDCT triage. The joint study of COVID-19 vaccine effectiveness in St. Petersburg was registered at ClinicalTrials.gov (NCT04981405, date of registration: August 4, 2021). This publication covers the data collected at the two outpatient centres of the MIBS that contributed to the study data. The Research Planning Board of the European University at St. Petersburg and the Ethics Committee of the Clinic Scandinavia approved the seroprevalence study on May 20, 2020 and May 26, 2020, respectively. Consent was obtained from all participants of the study. The study was registered with the following identifiers: Clinicaltrials.gov (NCT04406038, submitted on May 26, 2020, date of registration: May 28, 2020) and ISRCTN registry (ISRCTN11060415, submitted on May 26, 2020, date of registration: May 28, 2020).

### Summary of Updates

Database for the study updated, few cases added.

